# Closing the Immunization Gap: Complete basic childhood vaccination and associated factors among children aged 12–23 months in Tanzania; a multilevel analysis

**DOI:** 10.1101/2025.06.03.25328870

**Authors:** Erick Donard Oguma, Elihuruma Eliufoo Stephano, Sahnun Ally Kessy, Jovin R. Tibenderana, Victoria Godfrey Majengo, Tegemea Patrick Mwalingo, Mussa Hassan Bago, Immaculata P. Kessy, Azan Abubakar Nyundo, Mtoro J. Mtoro

## Abstract

**Background:** Globally, vaccination prevents an estimated 4 to 5 million deaths annually, including hundreds of thousands of deaths among children under five. Despite significant global progress in childhood immunization, complete vaccination coverage remains low in Sub-Saharan Africa, including Tanzania. Therefore, we aimed to examine the coverage of complete basic childhood vaccinations and associated factors among children aged 12–23 months in Tanzania.

**Methods:** Analytical cross-sectional study of the 2022 Tanzania Demographic and Health Surveys data was conducted. The sampling frame was stratified by geographic region and urban/rural areas, using a two-stage sampling method that selected primary sampling units based on census enumeration areas, followed by household selection using probability systematic sampling. Multilevel logistic regression, accounting for the complex survey design, was used to identify individual and community-level factors associated with complete childhood vaccination. Adjusted odds ratios (OR) and 95% Confidence intervals (CI) were used to estimate the strength of association.

**Results:** The prevalence of complete basic childhood vaccination was 52.5% (95%CI: 49.6 - 55.3). Among these children, coverage for individual vaccines was high, with 86.5% receiving the first dose of measles and 91.0% receiving BCG. However, the percentage receiving the full dose was lower for polio (58.9%) and 90.0% for DPT. Mothers with primary, secondary/higher education, in middle wealth quintile, had more than four ANC visits, had 1-2 under-five children were more likely to completely vaccinate their children. At the community level, mothers in western, central zones and Zanzibar were more likely to vaccinate their children.

**Conclusion:** Complete basic childhood vaccination coverage in Tanzania was suboptimal and associated with various factors including maternal education, middle wealth, more ANC visits, and fewer young children exhibited higher vaccination odds. Western, central zones, and Zanzibar showed higher coverage. Targeted interventions addressing education, wealth, ANC, family size, and regional disparities would be crucial to improve vaccination rates in Tanzania.

## Background

Immunization is one of the most cost-effective public health interventions, each year preventing an estimated 4.4 million deaths (1). Despite ongoing efforts towards recovery and strengthening beyond pre-pandemic (2019) levels, a staggering 21 million children remained either unvaccinated or under-vaccinated in 2023 (2,3). The number of children who did not receive any vaccines, often referred to as zero-dose children, reached 14.5 million in 2019 (4).

Incomplete childhood vaccination has been shown to raise the risk of acquiring and transmitting infections that may be prevented, which can cause severe illness, complications, and even death (5–7). Furthermore, it may reduce herd immunity, which would facilitate the spread of infectious illnesses throughout the community (6,8).

Measles, polio, and diphtheria outbreaks are still a threat in many areas, and the World Health Organization (WHO) continues to emphasize the significance of attaining high immunization coverage to stop them. The WHO supports regional and national plans to accomplish this through frameworks such as the Measles and Rubella Strategic Framework and Immunization Agenda 2030 (9,10).

Despite the proven benefits of vaccination, coverage is still unequal, especially in developing countries where variations in uptake and access can pose serious public health issues (11,12). In Africa, about one out of five children does not receive all of the necessary and basic vaccines. Consequently, each year, vaccine-preventable diseases (VPDs) continue to affect over 30 million children under five in Africa. Of these, over half a million of them die each year, which represents 58% of all VPD-related deaths worldwide (13). The coverage of basic childhood vaccinations is low (59.4%) in Sub-Saharan Africa, with variation among the countries (14). Complete basic childhood immunization is still a significant public health concern in East Africa, where it is low (69.21% in 2016), ranging from 39.5% in Ethiopia to 85% in Burundi (15).

According to Tanzania Demographic Health Survey (2022), 53% of children age 12–23 months are fully vaccinated against all basic antigens, which is a decline from 75% in 2015-16 (Received one dose each of BCG and measles vaccines, and three doses each of polio and pentavalent (Diphtheria, Tetanus, Pertussis, Haemophilus influenzae, and Hepatitis B) (16). However, 23% of children aged 12–23 months are fully vaccinated according to the national schedule. Across regions, vaccination coverage ranges from 3% in Shinyanga to 66% in Kilimanjaro (16).

To improve immunization rates and decrease health disparities, public health policies must take into account the current state of basic childhood vaccination coverage and its associated factors. Therefore, this multilevel analysis aims to evaluate the coverage of complete basic vaccination and its associated factors among children aged 12 to 23 months in Tanzania using the 2022 Demographic and Health Survey. The study seeks to provide valuable insights for policymakers and health practitioners that will help to strengthen immunization efforts in Tanzania.

## Methods

### Data source and design

An analytical cross-sectional study was conducted using the secondary data from the 2022 TDHS. The TDHS conducts nationally representative household surveys every five years. The survey was conducted by the Tanzania National Bureau of Statistics, collaborating with the Ministries of Tanzania Mainland and Zanzibar.

### Population and sampling

Data for this study were obtained from the recent DHS conducted between 24 February and 21 July 2022 across all regions in Tanzania. The target population for the 2022 TDHS included women of reproductive age (15–49 years) across the 32 administrative regions in Tanzania. At the country level, a sampling frame is usually obtained. To minimize sampling errors, the country is stratified by geographic region and by urban/rural areas within each region, followed by a two-stage sampling to select a household to be surveyed. The first sampling is to select a primary sampling unit (PSU) and then select a household. PSUs are survey clusters that are usually based on census enumeration areas (EAs). A probability proportional to size is employed in each stratum to select the PSU. For each selected PSU, a complete household listing is done. This is followed by selecting a fixed number of households to be surveyed using equal probability systematic sampling. Our analysis utilized the Kids Record file (KR), which initially contained 10,783 records. We excluded children who had died, those under 12 months of age, and those aged ≥24 months, resulting in a final weighted sample of 2,180 children.

### Study Variables

#### Outcome variable

The dependent variable was complete basic childhood vaccination status among children aged 12–23 months. Following WHO recommendations, this included receiving one dose each of BCG and measles vaccines, and three doses each of polio and pentavalent (diphtheria, tetanus, pertussis, Haemophilus influenzae, and hepatitis B) vaccines by the age of 12 months. Children who received all these recommended doses were categorized as “yes” (fully vaccinated), while those who did not were categorized as “no” (not fully vaccinated). Information on child vaccination was obtained from mothers’ verbal reports and data extracted from childhood immunization cards.

#### Explanatory variables

We included explanatory variables based on available dataset and literature; At the individual level we included maternal characteristics; age in years (15-24, 25-34 or 35-49), marital status (never married, married/cohabiting and previously married), education level (no formal education, primary education and secondary/higher), literacy (literate or illiterate), wealth index (poorest, poorer, middle, richer and richest), media exposure ((yes as listening to the radio, reading the newspaper or watching Television less than once a week or at least once a week or no if otherwise), working status (working or not working), sex of household head (male or female), household members (<6 or ≥6), ANC visits (<4 or ≥4), place of delivery (home or health facility), parity (primiparous, multiparous and grand multiparous), PNC checkup (yes or now), distance to the facility (big problem or nor a big problem). Children’s related characteristics included child sex (male or female), birth order (1st, 2nd & 3rd, or 4th & more), and number of under-five children (none, 1-2, and ≥3).

Community variables include residence (urban or rural), geographical zones (western, northern, central, southern highlands, southern, southwest highlands, lake, eastern and Zanzibar). Community literacy was calculated based on the proportion of women in each cluster based on literacy category, classified as either low (communities where <50% of women are literate) or high (communities where ≥50% of women are literate).

#### Data management and analysis

To account for the complexity of the survey design, we applied individual sampling weights, primary sampling units (clusters), and strata to adjust for the cluster sampling design. All analyses were performed using STATA version 18.5 (STATA Corp, College Station, TX). Descriptive statistics were summarized using means, standard deviation (SD), medians, frequency, and proportion for categorical variables. The Pearson chi-square test was used to compare the differences in the proportion of vaccination status across participants’ characteristics. Given the hierarchical structure of the DHS data, where women are nested within households and households are nested within clusters, there is likely greater similarity among women within the same cluster. This results in a violation of the assumption of independent observations and equal variance across clusters. Therefore, we used multilevel mixed-effects logistic regression. Four models were applied: the null model (outcome variable only), Model I (only individual-level factors), Model II (only community-level factors), and Model III (both individual and community-level factors.

Random effects were used to estimate the mean distribution of effects and compare individuals from two randomly chosen clusters. Intra-class correlation coefficient (ICC), Median odds ratio (MOR), and Proportional Change in Variance (PCV) (17–19). The ICC was calculated to assess the magnitude of the clustering effect and to evaluate the extent to which community-level factors account for the unexplained variance in the null model. When randomly picking out two clusters, the MOR was defined as the median value of the odds ratio between the area at the highest risk and the area at the lowest risk. The PCV was also computed for each model with respect to the null model to show the influence the cluster variables have on complete childhood vaccination. The PCV was calculated by PCV=(V_e_–V_mi_)/V_e_, where V_e_ is the variance in the null model and V_mi_ is the variance in successive models. The model that best suited the data was the one with the lowest deviance (Model III). Adjusted Odds ratio (AOR) and corresponding 95% confidence intervals (CI) were presented to estimate the magnitude and strength of the association. We compared the modified Poisson regression model and the logistic regression model using the Akaike Information Criterion (AIC), as the odds ratio might overestimate the association when the proportion of outcome is not rare (> 10%). Despite this, the logistic regression model was chosen because it had the lowest AIC, suggesting a better fit. A statistically significant result was considered for a p-value < 0.05. A variance inflation factor (VIF) was used to assess for multicollinearity between independent variables before fitting a multivariable regression model.

## RESULTS

### Sociodemographic characteristics of study participants

The mean age of mothers who had their child vaccinated was 28.7 years (standard deviation=6.9), with 44.0% aged 25-34 years. The majority (83.6%) were married or cohabiting, and just 7.0% were never married. More than half (54.9%) had completed primary education, and 71.6% were literate. Regarding socioeconomic status, 23.6% were from the poorest and 18.9% from the richest households. Nearly two-thirds (60.5%) were working, and 62.9% were exposed to media. Most (81.6%) were delivered at the health facility, and 62.3% attended more than four ANC visits. More than half of the children were male, and 21.9% were firstborn. More than half (72.4%) were from a rural setting, and 33.8% from the lake zone. There was a significant difference in individual and community level characteristics (p<0.05) with education, literacy, wealth index, media exposure, ANC visits, place of delivery, and number of under-five children. (Table 1).

**Table 1:** Individual and community level characteristics and distribution of vaccination status among children aged 12-23 months in Tanzania (N=2,180)

| Characteristics | N (%) | Vaccination status, N (%) |  | p-value |
| --- | --- | --- | --- | --- |
|  |  | Partial | Complete |  |
| **Mother's age (years)** |  |  |  | 0.769 |
| 15-24 | 732 (33.6) | 355 (48.5) | 377 (51.5) |  |
| 25-34 | 960 (44.0) | 457 (47.6) | 503 (52.4) |  |
| 35-49 | 488 (22.4) | 224 (45.9) | 264 (54.1) |  |
| Mean (±SD) | | | | |
| **Marital Status** |  |  |  | 0.137 |
| Never married | 154 (7.0) | 62 (40.0) | 92 (60.0) |  |
| Married/Cohabiting | 1822 (83.6) | 868 (47.6) | 954 (52.4) |  |
| Previously married | 205 (9.4) | 107 (52.1) | 98 (47.9) |  |
| **Education Level** |  |  |  | <0.001 |
| No formal education | 483 (22.2) | 275 (56.9) | 208 (43.1) |  |
| Primary | 1197 (54.9) | 561 (46.9) | 636 (53.1) |  |
| Secondary/Higher | 501 (23.0) | 200 (40.0) | 301 (60.0) |  |
| **Literacy** |  |  |  | 0.001 |
| Illiterate | 620 (28.4) | 344 (55.5) | 276 (44.5) |  |
| Literate | 1561 (71.6) | 692 (44.3) | 869 (55.7) |  |
| **Wealth index** |  |  |  | 0.004 |
| Poorest | 514 (23.6) | 296 (57.5) | 218 (42.5) |  |
| Poorer | 438 (20.1) | 196 (44.8) | 242 (55.2) |  |
| Middle | 392 (18.0) | 170 (43.3) | 222 (56.7) |  |
| Richer | 424 (19.4) | 186 (43.9) | 238 (56.1) |  |
| Richest | 413 (18.9) | 188 (45.6) | 225 (54.4) |  |
| **Media Exposure** |  |  |  | 0.008 |
| No | 810 (37.1) | 420 (51.8) | 390 (48.2) |  |
| Yes | 1371 (62.9) | 616 (45.0) | 755 (55.0) |  |
| **Working Status** |  |  |  | 0.601 |
| Not working | 861 (39.5) | 417 (48.5) | 444 (51.5) |  |
| Working | 1319 (60.5) | 618 (46.9) | 701 (53.1) |  |
| **Sex of Household Head** |  |  |  | 0.721 |
| Male | 1744 (80) | 833 (47.7) | 911 (52.3) |  |
| Female | 437 (20) | 203 (46.6) | 233 (53.4) |  |
| **Household members** |  |  |  | 0.175 |
| <6 | 1021 (46.8) | 464 (45.5) | 557 (54.5) |  |
| ≥6 | 1159 (53.2) | 572 (49.3) | 588 (50.7) |  |
| **ANC Visits** |  |  |  | 0.012 |
| <4 | 821 (37.7) | 423 (51.5) | 398 (48.5) |  |
| ≥4 | 1359 (62.3) | 613 (45.1) | 746 (54.9) |  |
| **Place of delivery** |  |  |  | 0.012 |
| Home | 401 (18.4) | 222 (55.3) | 179 (44.7) |  |
| Health facility | 1779 (81.6) | 814 (45.7) | 965 (54.3) |  |
| **Parity** |  |  |  |  |
| Primiparous | 445 (20.4) | 205 (46.1) | 240 (53.9) | 0.854 |
| Multiparous | 1162 (53.3) | 554 (47.7) | 608 (52.3) |  |
| Grand multiparous | 574 (26.3) | 277 (48.3) | 297 (51.7) |  |
| **PNC check-up** |  |  |  | 0.356 |
| **(n=1,675)** |  |  |  |  |
| No | 635 (37.9) | 278 (43.8) | 357 (56.2) |  |
| Yes | 1040 (62.1) | 488 (46.9) | 552 (53.1) |  |
| **Child sex** |  |  |  | 0.451 |
| Male | 1139 (52.3) | 530 (46.5) | 610 (53.5) |  |
| Female | 1041 (47.7) | 506 (48.6) | 535 (51.4) |  |
| **Birth order** |  |  |  | 0.896 |
| 1st | 477 (21.9) | 223 (46.7) | 254 (53.3) |  |
| 2nd & 3rd | 873 (40.0) | 412 (47.2) | 461 (52.8) |  |
| 4th & more | 831 (38.1) | 401 (48.3) | 430 (51.7) |  |
| **Number of under-five children** |  |  |  | 0.041 |
| None | 48 (2.2) | 27 (57.6) | 21 (42.4) |  |
| 1-2 | 1,736 (79.6) | 796 (45.9) | 940 (54.1) |  |
| ≥3 | 397 (18.2) | 212 (53.5) | 185 (46.5) |  |
| **Distance to the facility** |  |  |  | 0.107 |
| Big problem | 734 (33.7) | 373 (50.8) | 361 (49.2) |  |
| Not a big problem | 1,446 (66.3) | 663 (45.8) | 783 (54.2) |  |
| **Residence** |  |  |  | 0.593 |
| Urban | 603 (27.6) | 279 (46.2) | 324 (53.8) |  |
| Rural | 1,578 (72.4) | 757 (48.0) | 820 (52.0) |  |
| **Geographical Zones** |  |  |  | 0.055 |
| Western | 230 (10.5) | 109 (47.6) | 121 (52.4) |  |
| Northern | 226 (10.4) | 105 (46.4) | 121 (53.6) |  |
| Central | 222 (10.2) | 92 (41.6) | 130 (58.4) |  |
| Southern highlands | 124 (5.7) | 45 (36.6) | 79 (63.4) |  |
| Southern | 84 (3.9) | 50 (59.4) | 34 (40.6) |  |
| Southwest highlands | 200 (9.2) | 108 (53.7) | 92 (46.3) |  |
| Lake | 737 (33.8) | 361 (49.0) | 376 (51.0) |  |
| Eastern | 298 (13.7) | 148 (46.0) | 150 (50.4) |  |
| Zanzibar | 61 (2.8) | 19 (30.3) | 42 (69.7) |  |

### Prevalence of complete basic vaccination

The overall prevalence of complete basic vaccination among children aged 12-23 months was 52.5% (95% CI: 49.6-55.3). Among these children, coverage for individual vaccines was high, with 86.5% receiving the first dose of measles and 91.0% receiving BCG. However, the percentage receiving the full series was lower for polio (58.9% for all three doses) and DPT (90.0% for all three doses) (Figure 1).

**Figure 1:**
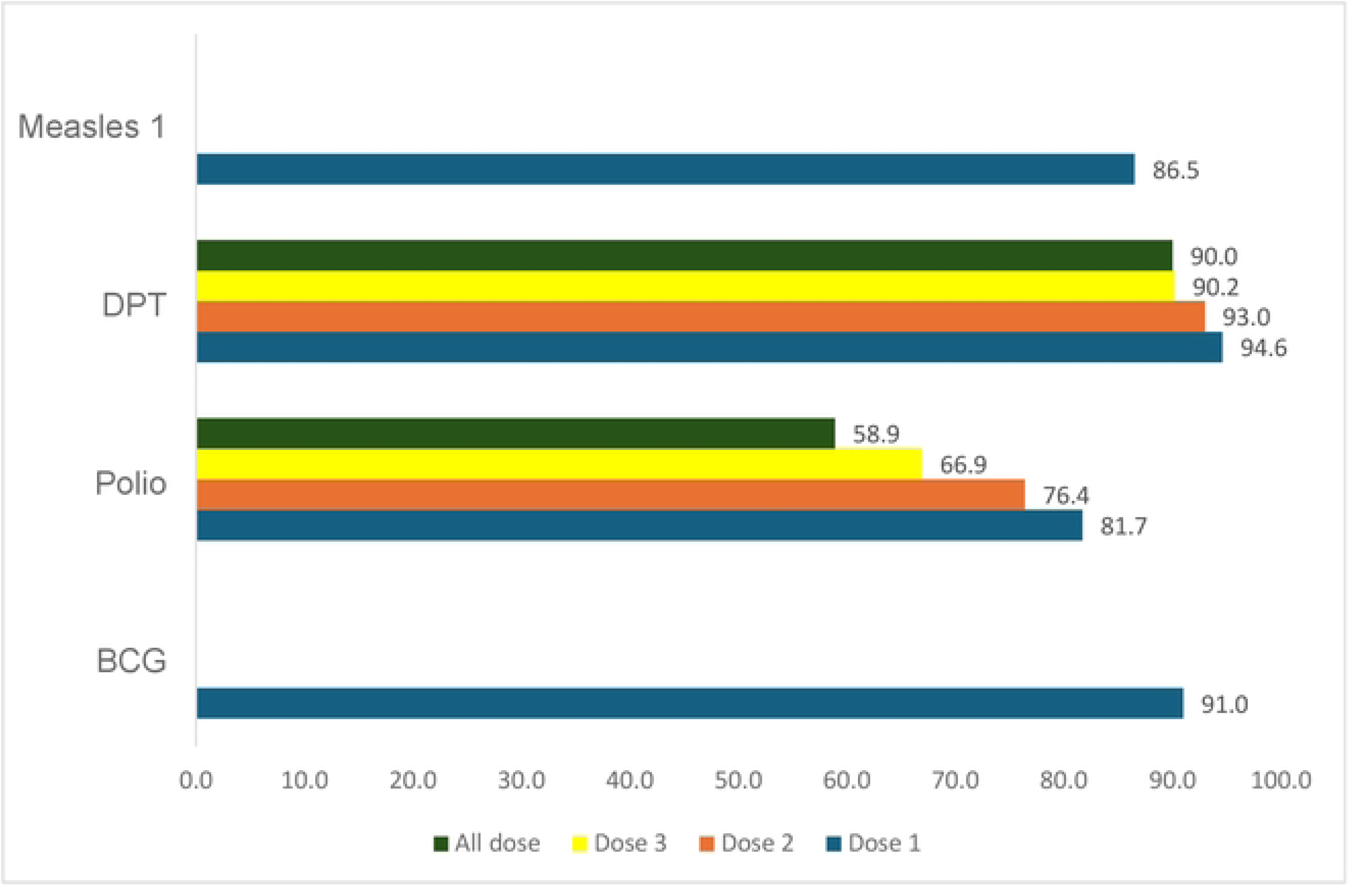
Distribution of basic vaccination among children aged 12-23 months in Tanzania.

### Factors associated with vaccination status

From the final multivariable analyses, mothers with primary education (AOR=1.51, 95%CI: 1.16-1.99) and secondary/higher (AOR=1.92, 95%CI: 1.35-2.74) were more likely to vaccinate their children than their counterparts. Mothers with more than four ANC visits were 25% more likely to attain complete vaccination than their counterparts (AOR=1.25, 95%CI:1.02-1.53). Households with 1-2 under-five children were more likely to vaccinate their children compared to those with no under-five children (AOR=2.23, 95%CI: 1.19-4.15). Geographically, mothers in the western zone (AOR=2.18, 95%CI: 1.13-4.22), central zone (AOR=2.13, 95%CI: 1.11-4.07), and Zanzibar (AOR=3.12, 95%CI: 1.64-5.92) had higher odds of vaccination of their children than those in the southern zone. (Table 2).

**Table 2:**
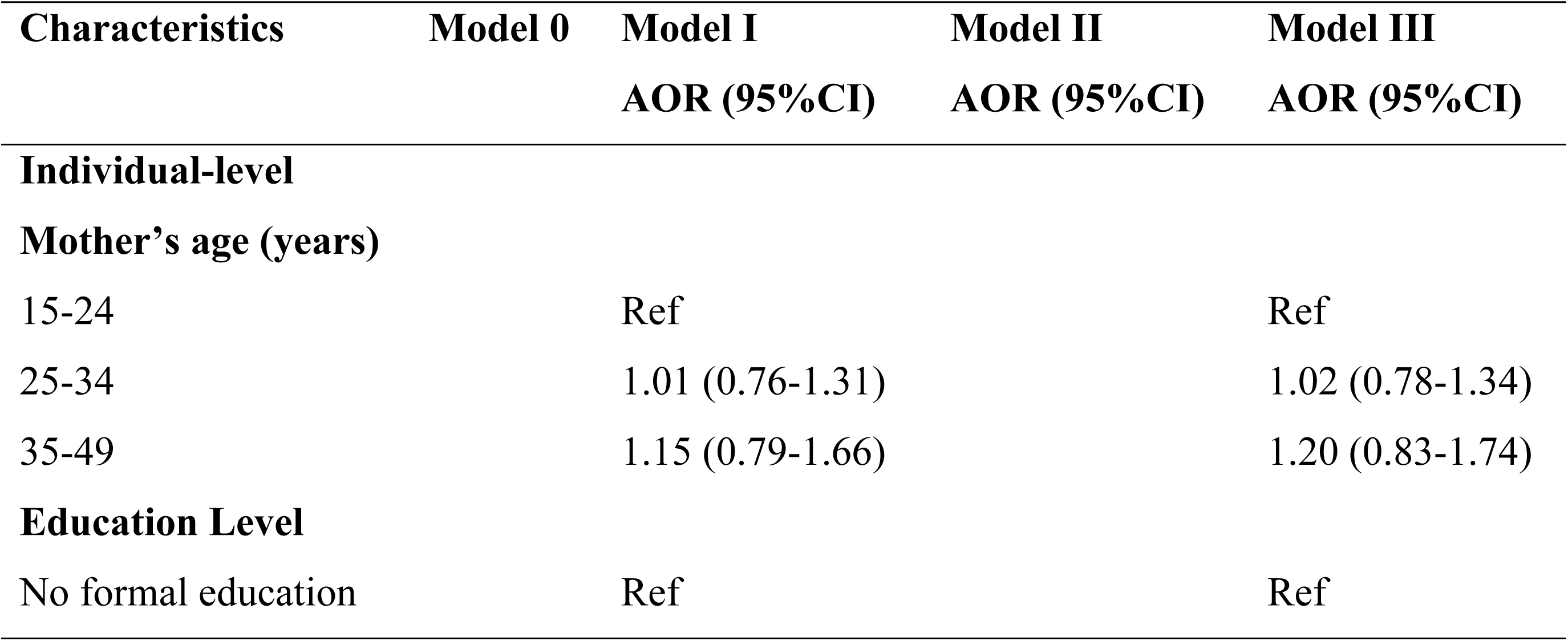
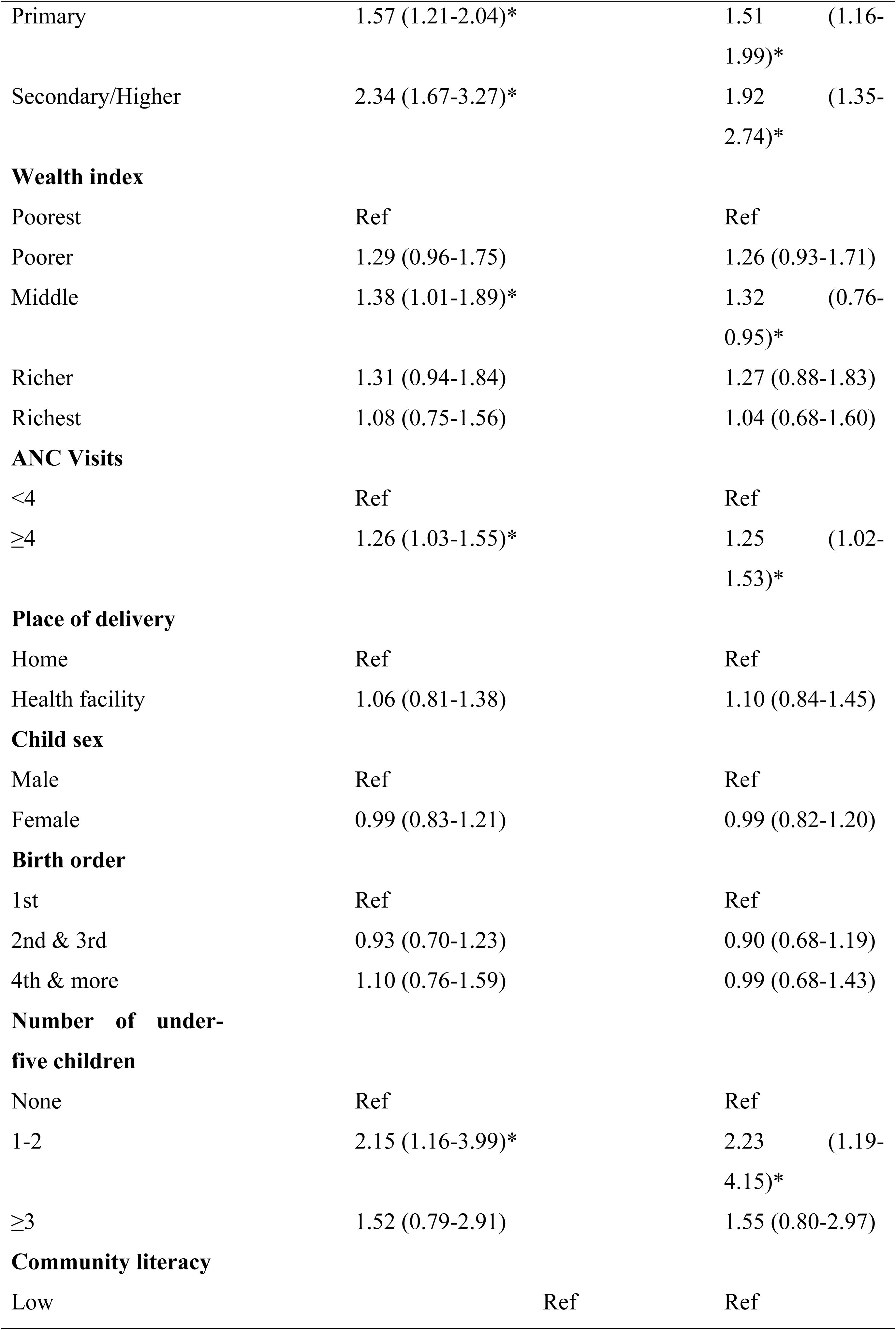
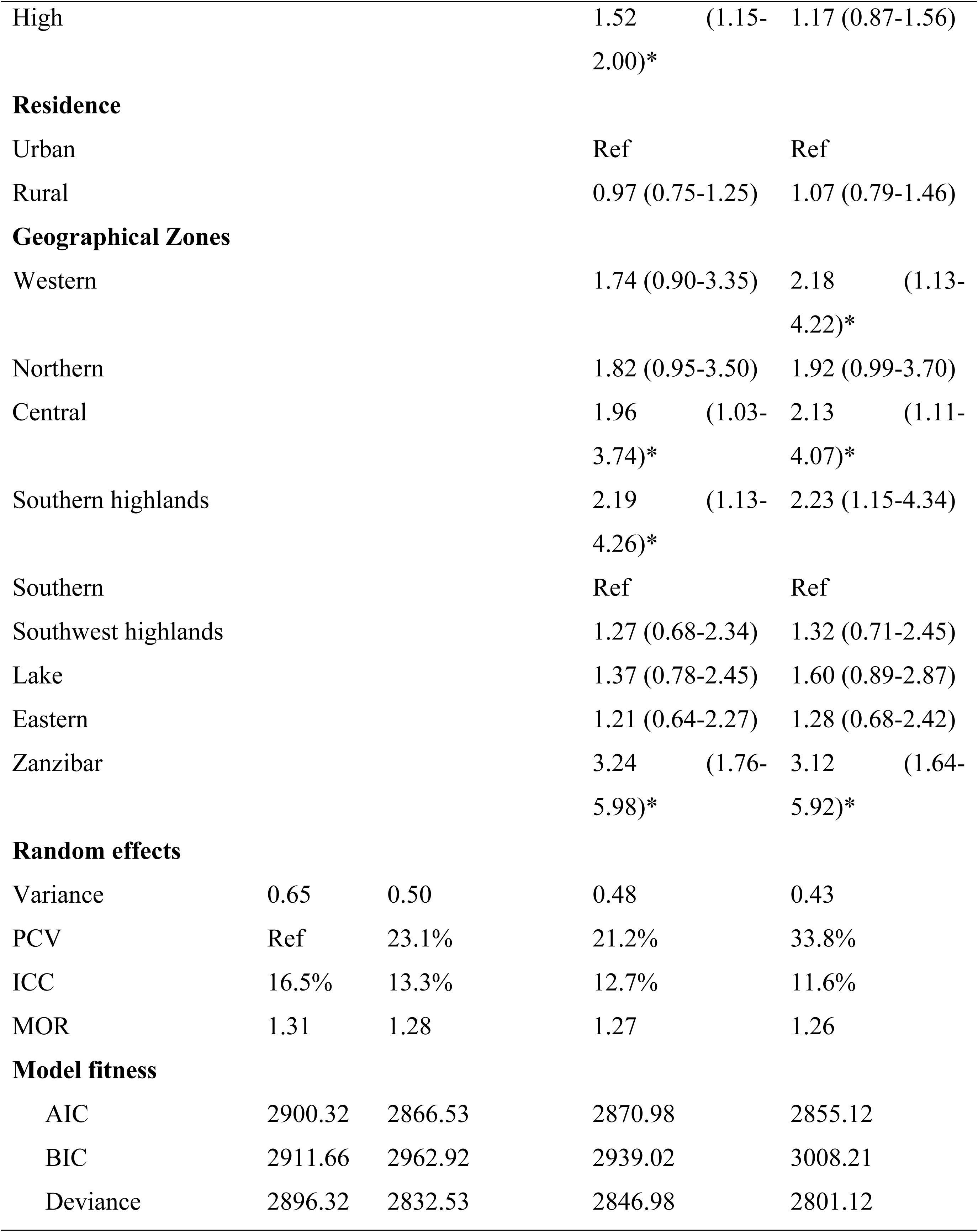
Individual and community factors associated with complete vaccination among children aged 12-23 months in Tanzania.

### Random effects and model fitness

The null model showed a variance of 0.65 and a p-value of < 0.001, indicating significant differences in complete vaccination across localities. In this model, the odds of complete vaccination varied by a factor of 1.31 (MOR). According to the Intraclass Correlation Coefficient (ICC) in Model I, 13.3% of the variation in complete vaccination was attributed to individual differences. In Model I, the likelihood of complete vaccination was 1.28 times greater than for incomplete vaccination. The best-fitting model was Model III, which had the lowest deviance (2801.12) and AIC (2855.12). (Table 2).

## Discussion

Childhood immunization remains a critical intervention for preventing and protecting children from preventable childhood infections (20). However, incomplete childhood vaccination remains a global concern in developing countries (7,13,21). This study intends to evaluate the coverage of complete basic vaccination and its associated factors among children aged 12 to 23 months in Tanzania. The study seeks to generate evidence-based recommendations that will aid in developing cost-effective public interventions and inform health policy.

The study revealed that more than fifty percent of children, 52.5%, had completed their basic vaccination. Full basic vaccination coverage among 12-23 months children in this study reveals a substantial decline of 22.5% from 75% in the previous Survey 2015-16 (16). The possible reason for this sudden drop may be influenced by COVID-19 disruption, an increase in population, geographical access barriers, and rural-urban disparities.

Furthermore, inconsistency in complete basic vaccination coverage was observed in previous studies conducted in Ethiopia, Malawi, and East Africa, these studies reported high prevalence of complete basic childhood vaccination, above 69% (15,22,23). The discrepancy in these findings might be attributed by their variation in sample size and study setting. Low prevalence of complete basic vaccination in this study create the need for immediate public intervention. As evidenced, incomplete childhood vaccination has been shown to raise the risk of acquiring infections that may be prevented, which can cause severe sickness, complications, and even death (5–7). Furthermore, incomplete vaccination schedule may reduce herd immunity, which would facilitate the spread of infectious illnesses throughout the community (6,8).

Moreover, the finding revealed that mothers with primary and secondary or higher were more likely to vaccinate their children completely than women with no education. These findings are consistent with those reported from the study done in Mozambique, Ethiopia, East Africa, and Ghana, which also reported high education to be associated with full vaccination among children (15,24–26).

The possible reason could be that mothers who were less educated or never attended school are more likely to be less informed or aware of the health benefits of childhood immunization and the negative consequences of incomplete childhood immunization. Additionally, low education has been reported to be associated with negative perception toward childhood immunization, poor child clinic attendance, and little child health attention, which further increases the chance of incomplete childhood vaccination (21,27).

Based on these results, the study underscores the importance of providing community education on childhood immunization while putting more effort into educating women on the benefits of full vaccination and the effects of incomplete childhood vaccination. This would be beneficial if women were informed early; therefore, effective integration of immunization education in antenatal care would be a good starting point.

Moreover, the study revealed that mothers who have attended four antenatal care visits and above are more likely to complete vaccinating their children than those who attend fewer than four visits. These results align with the findings reported from the previous study in Ethiopia, East Africa, and Ghana (15,28). The similarity in these studies could be explained by the fact that women who attend more than 4 ANC visits are more likely to be taught more health information (education) about their expected future baby, including immunization and breastfeeding. Hence, emphasizing that women to attend antenatal care is one of the key aspects in maximizing full vaccination coverage.

Similarly, the study revealed the wealth index as one of the important determinants of complete basic vaccination in children. The finding shows that mothers with a middle wealth index are more likely to fully vaccinate their children compared to mothers with a poor wealth index. This finding is supported by the study conducted in Malawi and East Africa (15,23).

Due to the cost incurred during transport, Poor economic status may limit a mother from accessing the health Centre, particularly women who live a far distance from the health facility (22,24,26). This may expose these women to a high risk of missing some vaccine. The situation will even be more severe for single-mother families and unmarried mothers who are the primary providers in their family (22,29). Because of financial insecurity, they are spending most of their time looking for money to take care of their family, which may minimize their attention on child health matters, such as taking a child to clinics. They may delegate this responsibility to the house girls most of the time.

This study underscores the importance of empowering women on major economic issues. This could be achieved by creating equal opportunity, minimizing barriers to access to economic activities, providing affordable capital for economic development, providing more economic activities, and strengthening gender mainstreaming.

Furthermore, the study revealed that households with 1-2 under-five children were more likely to fully vaccinate their children than those without under-five children. This finding correlates with the previous studies conducted in Somalia (30). The possible reason for this similarity is that women with 1-2 children were more likely to have vaccination experience than those without children or who had never been exposed to vaccination issues. Prior childhood immunization experience may influence their decision whether their child should be fully vaccinated. Additionally, these mothers are more likely to have more knowledge about childhood vaccination, which may affect their decision.

Additionally, the study shows that although there is a high initial vaccination uptake (e.g., 91% for BCG, 86.5% for measles), only 52.5% of children obtain all of the recommended basic immunizations, with notable declines for vaccines such as polio (58.9%). This emphasizes the need for policies that increase health education, especially for mothers with less education and fewer ANC visits, strengthen follow-up procedures, and address the challenges presented by lower-income and larger households. Regional differences also highlight the significance of duplicating effective models from high-coverage zones, such as the central region and Zanzibar, in order to adapt methods to underperforming areas. Boosting full vaccination coverage will require improving community participation, boosting access, and integrating immunization with mother and child health services.

### Strengths and limitations

This study’s strengths include using nationally representative 2022 TDHS data and a large sample, enhancing the generalizability of our findings to the Tanzanian population. The rigorous data collection, involving experienced field assistants, ensured high data quality and reliability. Moreover, multilevel binary logistic regression provided a robust analysis, effectively accounting for survey design complexities and strengthening the validity of our conclusions. However, the cross-sectional nature of the data prevents establishing a causal relationship.

### Implications for practice and policy recommendations

The suboptimal rate of complete basic childhood vaccination in Tanzania necessitates targeted and multi-faceted interventions. Given the positive influence of maternal education, healthcare providers should prioritize clear and accessible vaccination information during antenatal and postnatal care visits, tailoring messages to mothers with varying levels of education. Furthermore, the higher coverage observed in the western, central zones, and Zanzibar warrants investigation to identify and replicate successful local practices in other regions. To improve complete childhood vaccination coverage, policymakers should invest in and strengthen initiatives promoting girls’ education and women’s literacy nationwide. Targeted financial mechanisms would be essential to ensure equitable access to vaccination services for vulnerable populations. Integrating comprehensive immunization education into enhanced antenatal care services is essential.

## Conclusion

Complete basic childhood vaccination coverage in Tanzania was suboptimal and associated with various factors, including maternal education, middle wealth, more ANC visits, and fewer young children exhibiting higher vaccination odds. Western, central zones, and Zanzibar showed higher coverage. Targeted interventions addressing education, wealth, ANC, family size, and regional disparities would be crucial to improving vaccination rates in Tanzania.

## Data Availability

DHS data are publicly available at https://dhsprogram.com, but the dataset supporting the conclusion is available with the corresponding author and can be shared upon a reasonable request.

## Abbreviations

AIC: Akaike Information Criterion
ANC: Antenatal Care
AOR: Adjusted odds ratio
BCG: Bacillus Calmette-Guérin
BIC: Bayesian Information Criterion
DHS: Demographics and Health Survey
DPT: Diphtheria-Pertussis-Tetanus
EAs: Enumeration areas
ICC: Intra-class Correlation Coefficient
LLR: Likelihood ratio
MOR: Median Odds Ratio
PCV: Proportional Change in Variance
PSU: Primary sampling unit
SSA: Sub-Saharan Africa
TDHS: Tanzania Demographics and Health Survey
VIF: Variance inflation factor

## Acknowledgements

We thank the DHS program for making the data available for this study and TILAM International for statistical consultation.

## Declarations

### Ethics approval and consent to participate

This study utilized publicly available, de-identified data from the 2022 TDHS, accessible online through the DHS program. The original survey received ethical approval from both the National Institute of Medical Research Ethics Committee in Tanzania and the ICF Macro Ethics Committee in Calverton, New York. Permission to use the data for this secondary analysis was granted by the DHS program upon acceptance of the proposed analysis plan under the designated account, with credentials available upon request via https://dhsprogram.com/data/dataset_admin/index.cfm. As this study involved secondary data analysis of publicly accessible datasets, no additional ethical approval was required. Informed consent was obtained from all participants during the initial survey, and all procedures adhered strictly to relevant guidelines and regulations. Further details regarding DHS data usage and ethical standards can be found at http://goo.gl/ny8T6X.

### Consent for publication

Not applicable.

## Reference

1. Carter A, Msemburi W, Sim SY, Gaythorpe KAM, Lambach P, Lindstrand A, et al. Modeling the impact of vaccination for the immunization Agenda 2030: Deaths averted due to vaccination against 14 pathogens in 194 countries from 2021 to 2030. Vaccine. 2024;

2. Carter A, Msemburi W, Sim SY, A.M. Gaythorpe K, Lindstrand A, Hutubessy RCW. Modeling the Impact of Vaccination for the Immunization Agenda 2030: Deaths Averted Due to Vaccination Against 14 Pathogens in 194 Countries from 2021-2030. SSRN Electron J [Internet]. 2021 Apr 27 [cited 2025 May 25]; Available from: https://data.unicef.org/topic/child-health/immunization/

3. UNICEF. Vaccination and Immunization Statistics - UNICEF DATA [Internet]. [cited 2025 May 25]. Available from: https://data.unicef.org/topic/child-health/immunization/

4. World Health Organization (WHO). Immunization coverage [Internet]. [cited 2025 May 25]. Available from: https://www.who.int/news-room/fact-sheets/detail/immunization-coverage

5. Kanchanarat S, Chinviriyasit S, Chinviriyasit W. Mathematical Assessment of the Impact of the Imperfect Vaccination on Diphtheria Transmission Dynamics. Symmetry (Basel). 2022;

6. Magpantay FMG, Riolo MA, Domenech De Cèlles M, King AA, Rohani P. Epidemiological consequences of imperfect vaccines for immunizing infections. SIAM J Appl Math. 2014;

7. Atnafu Gebeyehu N, Abebe Gelaw K, Asmare Adella G, Dagnaw Tegegne K, Adie Admass B, Mesele Gesese M. Incomplete immunization and its determinants among children in Africa: Systematic review and meta-analysis. Hum Vaccines Immunother. 2023;

8. Rashid H, Khandaker G, Booy R. Vaccination and herd immunity: What more do we know? Current Opinion in Infectious Diseases. 2012.

9. World Health Organization. Immunization agenda 2030. Who [Internet]. 2022;1–58. Available from: https://www.who.int/immunization/ia2030_Draft_One_English.pdf?ua=1

10. World Health Organization. World Health Organization. Measles and rubella strategic framework 2021-2030 [Internet]. 2024. Available from: https://www.who.int/publications/i/item/measles-and-rubella-strategic-framework-2021-2030

11. Bobo FT, Asante A, Woldie M, Dawson A, Hayen A. Child vaccination in sub-Saharan Africa: Increasing coverage addresses inequalities. Vaccine. 2022;

12. Lai X, Zhang H, Pouwels KB, Patenaude B, Jit M, Fang H. Estimating global and regional between-country inequality in routine childhood vaccine coverage in 195 countries and territories from 2019 to 2021: a longitudinal study. eClinicalMedicine. 2023;

13. World Health Organization. Business case for WHO immunization activities on the African continent 2018-2030. 2018. 40 p.

14. Fenta SM, Biresaw HB, Fentaw KD, Gebremichael SG. Determinants of full childhood immunization among children aged 12–23 months in sub-Saharan Africa: a multilevel analysis using Demographic and Health Survey Data. Trop Med Health. 2021;

15. Tesema GA, Tessema ZT, Tamirat KS, Teshale AB. Complete basic childhood vaccination and associated factors among children aged 12–23 months in East Africa: a multilevel analysis of recent demographic and health surveys. BMC Public Health. 2020;

16. Ministry of Health (MoH) [Tanzania Mainland], Ministry of Health (MoH) [Zanzibar], National Bureau of Statistics (NBS), Office of the Chief Government Statistician (OCGS), and ICFMinistry of Health (MoH) [Tanzania Mainland], Ministry of Health (MoH) [Zanz and I. Tanzania Demographic and Health Survey and Malaria Indicator Survey 2022 Key Indicators Report. 2023;1–23.

17. Austin PC, Merlo J. Intermediate and advanced topics in multilevel logistic regression analysis. Stat Med. 2017;36(20):3257–77.

18. Merlo J, Wagner P, Ghith N, Leckie G. An original stepwise multilevel logistic regression analysis of discriminatory accuracy: The case of neighbourhoods and health. PLoS One. 2016;11(4):1–31.

19. Dong N, Reinke WM, Herman KC, Bradshaw CP, Murray DW. Meaningful Effect Sizes, Intraclass Correlations, and Proportions of Variance Explained by Covariates for Planning Two- and Three-Level Cluster Randomized Trials of Social and Behavioral Outcomes. Eval Rev. 2016;40(4):334–77.

20. World Health Organization (WHO). Immunization | WHO | Regional Office for Africa [Internet]. [cited 2025 May 25]. Available from: https://www.afro.who.int/health-topics/immunization

21. Bangura JB, Xiao S, Qiu D, Ouyang F, Chen L. Barriers to childhood immunization in sub-Saharan Africa: A systematic review. BMC Public Health. 2020.

22. Mekonnen AG, Bayleyegn AD, Ayele ET. Immunization coverage of 12-23 months old children and its associated factors in Minjar-Shenkora district, Ethiopia: A community-based study. BMC Pediatr. 2019;

23. Ntenda PAM. Factors associated with non- and under-vaccination among children aged 12–23 months in Malawi. A multinomial analysis of the population-based sample. Pediatr Neonatol. 2019;

24. Jani J V., De Schacht C, Jani I V., Bjune G. Risk factors for incomplete vaccination and missed opportunity for immunization in rural Mozambique. BMC Public Health. 2008;

25. Tesfa GA, Yehualashet DE, Getnet A, Bimer KB, Seboka BT. Spatial distribution of complete basic childhood vaccination and associated factors among children aged 12– 23 months in Ethiopia. A spatial and multilevel analysis. PLoS One. 2023;

26. Akanpaabadai EA, Adiak AA, Nukpezah RN, Adokiya MN, Adjei SE, Boah M. Population-based cross-sectional study of factors influencing full vaccination status of children aged 12-23 months in a rural district of the Upper East Region, Ghana. BMC Pediatr. 2024;

27. Vasudevan L, Baumgartner JN, Moses S, Ngadaya E, Mfinanga SG, Ostermann J. Parental concerns and uptake of childhood vaccines in rural Tanzania – a mixed methods study. BMC Public Health. 2020;

28. Kuuyi A, Kogi R. Factors contributing to immunization coverage among children less than 5 years in Nadowli-Kaleo District of Upper West Region, Ghana. PLOS Glob Public Heal [Internet]. 2024;4(8 August):1–15. Available from: 10.1371/journal.pgph.0002881

29. Kuroda H, Goto A, Kawakami C, Yamamoto K, Ito S, Kamijima M, et al. Association between a single mother family and childhood undervaccination, and mediating effect of household income: a nationwide, prospective birth cohort from the Japan Environment and Children’s Study (JECS). BMC Public Health. 2022;

30. Belay DB, Ali MI, Chen DG, Jama UA. Prevalence and associated factors of immunization among under-five children in Somalia. BMC Public Health [Internet]. 2025;25(1). Available from: 10.1186/s12889-025-22122-7

